# Cerebrovascular Response to an Acute Bout of Short Interval High Intensity Interval Exercise and Recovery in Healthy Adults

**DOI:** 10.1101/2021.05.05.21256704

**Authors:** Alicen A. Whitaker, Stacey E. Aaron, Carolyn S. Kaufman, Brady K. Kurtz, Stephen X. Bai, Eric D. Vidoni, Sandra A. Billinger

## Abstract

**Introduction:** High intensity interval exercise (HIIE) is performed widely. However, the field possesses limited knowledge regarding the acute HIIE cerebrovascular response. Our objective was to characterize the middle cerebral artery blood velocity (MCAv) response during an acute bout of short interval HIIE in healthy adults. We hypothesized MCAv would decrease below BL 1) during HIIE, 2) following HIIE, 3) and 30-minutes after HIIE. As a secondary objective, we investigated sex differences in the MCAv response during HIIE.

**Methods:** Fourteen healthy adults (7 male) completed the HIIE session. The 10-minute HIIE session included alternating 1-minute bouts of high-intensity and low-intensity intervals. MCAv, mean arterial pressure (MAP), heart rate (HR), and expired end tidal carbon dioxide (P_ET_CO_2_), were recorded at BL, during HIIE, following HIIE, and 30-minutes after HIIE.

**Results:** Contrary to our hypothesis, MCAv remained above BL for the HIIE duration. MCAv peaked at the third minute then decreased concomitantly with P_ET_CO_2_. MCAv was lower than BL after HIIE (p=0.03). Thirty minutes after HIIE, MCAv returned to near BL values (p = 0.47). Women showed higher BL MCAv (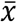 = 70.9 ± 8.1 cm·s^-1^) compared to men (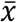 = 59.3 ± 5.8 cm·s^-1^, p = 0.01). A greater magnitude of MCAv response was observed in men resulting in non-significant differences during HIIE secondary to higher workload (p = 0.03).

**Conclusions:** Collectively, these findings show that in healthy adults, MCAv remained above BL during a 10-minute short-interval HIIE and returned to resting values 30 minutes after exercise.

## Introduction

Temporal changes in cerebral blood flow, measured by middle cerebral artery blood velocity (MCAv), have been well characterized during submaximal continuous exercise.(1–4) During steady-state low and moderate intensity continuous exercise, MCAv remains at a consistent rate.(2, 5) As exercise intensifies, MCAv increases in tandem up to anaerobic threshold, (1, 6, 7) thereafter MCAv decreases with high intensity exercise.(1, 4, 6) At the anaerobic threshold, hyperventilation occurs, resulting in decreased partial pressure carbon dioxide (PaCO_2_).(8) To attenuate the decrease in PaCO_2_, downstream arteriole vasoconstriction occurs causing a decrease in MCAv.(9)

While MCAv has been characterized during an acute bout of high intensity continuous exercise,(1, 3) our recent systematic review determined few studies have examined MCAv during an acute bout of high intensity interval exercise (HIIE).(10) HIIE is defined as repetitive short bouts of exercise switching between intervals of high intensity and low intensity recovery. In prepubertal children MCAv decreased below baseline (BL) during an acute 12-minute bout of HIIE with 1-minute intervals.(11) In contrast, healthy adults (average age of 23 years old) showed decreases in MCAv during an acute 20-minute bout of sprint interval exercise with repetitive 30-second sprint intervals at maximal effort with 4.5-minute active recovery. The authors report MCAv remained above BL throughout the duration of HIIE. The longer recovery interval of 4.5 minutes may have allowed for stabilization of PaCO_2_, therefore maintaining MCAv above baseline.(12)

Interval exercise at moderate intensity has been proposed to be beneficial for cerebrovascular function in older men due to the low intensity intervals allowing recovery of P_ET_CO_2_ and sustained MCAv.(13) However, sustained MCAv during HIIE has been stated to be counterintuitive due to decreases in expired end tidal carbon dioxide (P_ET_CO_2_).(13) Further, short interval HIIE with 1-minute low intensity intervals may not be enough time to allow for recovery of P_ET_CO_2_, leading to reductions of MCAv. Although this has been suggested, no studies have reported the effects of short interval HIIE on MCAv. To address this gap in knowledge, we proposed to characterize the MCAv response in healthy adults during a single bout of HIIE using 1-minute intervals of high and low intensity.

Our primary hypothesis was that MCAv would significantly decrease below BL MCAv during an acute 10-minute HIIE bout. Our secondary hypotheses were that MCAv would be significantly lower than BL MCAv 1) during passive recovery after HIIE and 2) at rest 30-minutes after HIIE. Further, we hypothesize that the decrease in MCAv will be related to the change in P_ET_CO_2_. Our previous work identified unique sex-specific trajectories in resting MCAv over the lifespan(14) and to moderate-intensity continuous exercise.(15) Therefore, as a secondary objective, we investigated whether sex differences existed in the MCAv response to HIIE.

## Methods

Individuals were recruited from the surrounding community using flyers. Participants were enrolled in the study if they met the following inclusion criteria: 1) 18-30 years old and 2) low cardiac risk defined by the American College of Sports Medicine (ACSM).(16) The study and all experimental procedures were approved by the University of Kansas Medical Center Human Subjects Committee. This study is registered on clinical trials.gov (NCT04673994). Prior to the start of study procedures, participants provided written informed consent.

Fifteen healthy adults (7 women, 8 men) completed the study consisting of two visits to the laboratory. The second visit occurred 48 hours to 2 weeks after the initial visit. No adverse events occurred. Due to loss of TCD signal during HIIE, we excluded one male individual from data analysis. Three participants required rescheduling for study visit 2 due to University closure for weather. These individuals completed study visit 2 within one month of the initial study visit. All participants confirmed they refrained from caffeine for 8 hours,(17) vigorous exercise for 24 hours,(18) and alcohol for 24 hours.(19) All participants reported no change in health or physical activity levels between study visits. For women, visits were scheduled between days 1-7 of the menstrual cycle.(20) Study visits were scheduled between 2-5pm to maintain consistency and minimize circadian influence on cerebral blood velocity.(21) All participants were classified as having <50% carotid stenosis.(22) Six out of seven (86%)women reported using birth control medication. Participant characteristics are presented in **Table 1**.

**Table 1.**
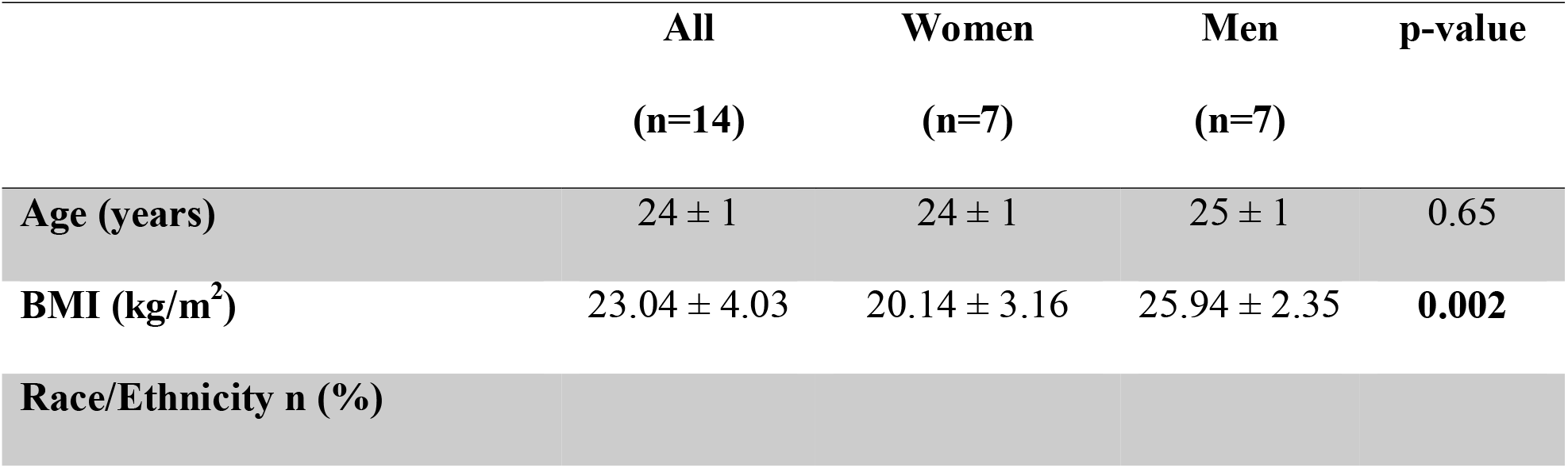
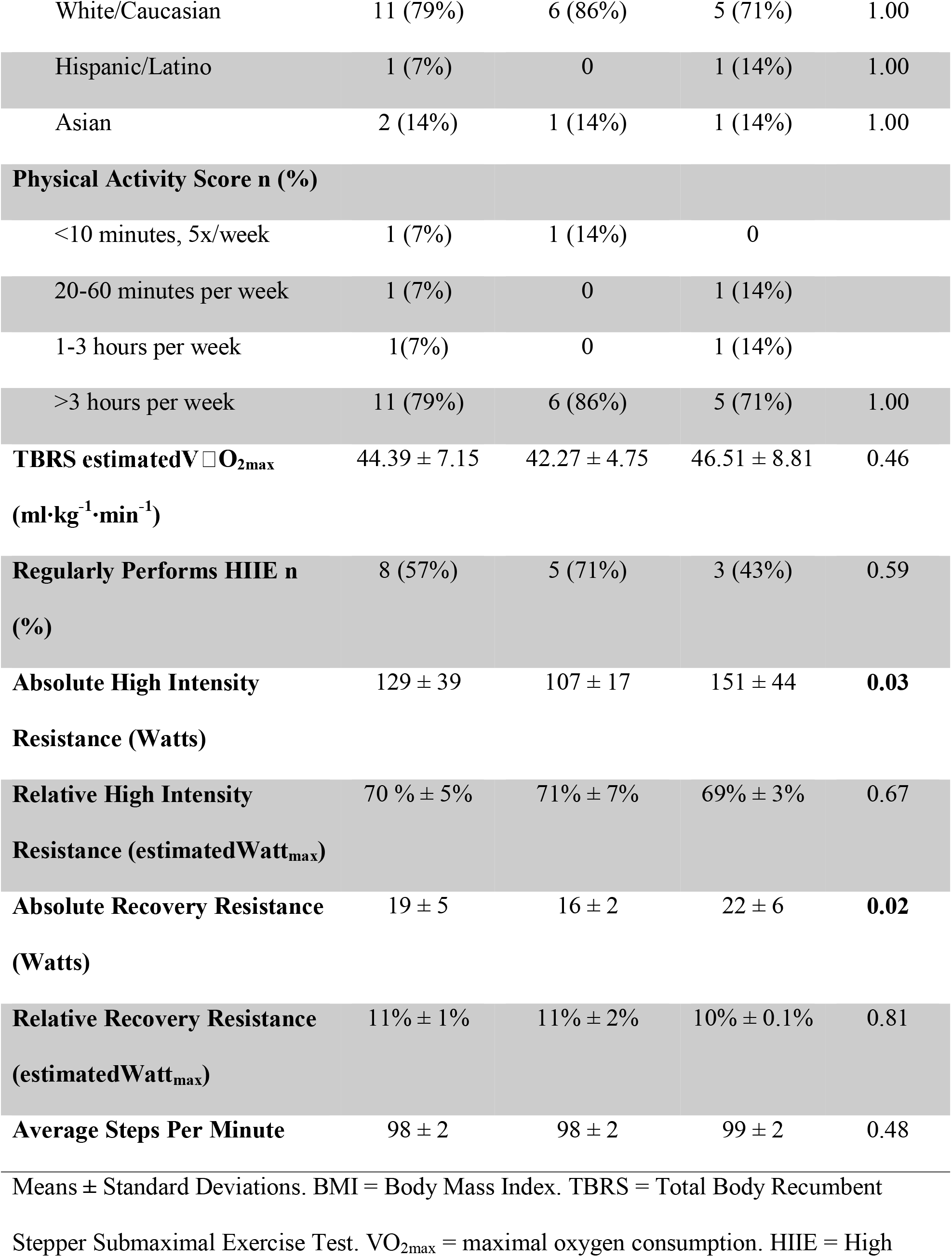
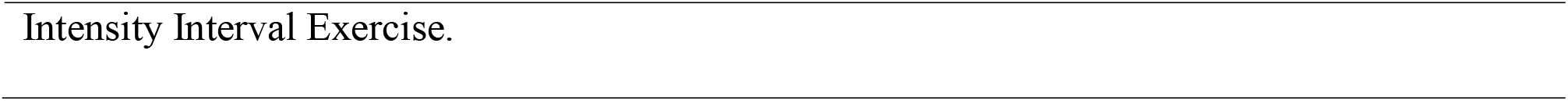
Participant Characteristics.

### Study Visit 1

We collected demographic information including age, sex, race/ethnicity, body mass index (BMI), self-reported physical activity, current medications, and medical history. Participants completed questionnaires including the ACSM Risk Stratification Screen.(23)

#### TCD Signal Acquisition

Following the questionnaires, participants were screened to ensure a MCAv signal on the right and/or left temporal window. Participants were fitted with an adjustable headband that secured bilateral transcranial Doppler ultrasound (TCD) probes (2-MHz, Multigon Industries Inc, Yonkers, New York) with ultrasonic gel onto the temporal region of the participant’s head. MCAv signal was acquired using standard probe positioning, orientation, depth, and direction.(24) The TCD depth, gain, power, and probe position were recorded to maintain consistency between study visits.

#### Submaximal Exercise Test

Participants performed the total body recumbent stepper (TBRS) submaximal exercise test(25, 26) to determine the estimated maximal workload.(13, 25, 26) Participants were fitted with a heart rate monitor (Polar Electro Inc, New York) and seated on a recumbent stepper (T5XR NuStep, Inc. Ann Arbor, MI). We performed the TBRS submaximal exercise test according to previously published methodology,(25) with the exception that participants were instructed to perform exercise by only using their lower extremities. For the HIIE acute exercise bout, participants would be exercising with their lower extremities since participants rested their arms on platforms to allow for accurate blood pressure measures. Therefore, we wanted to ensure an accurate workload for exercise prescription. All participants began exercising at 30 watts and stepped at a constant pace between 95-100 steps per minute. Every minute, heart rate (HR) and the rating of perceived exertion (RPE) were measured until termination criteria were met: 1) completion of all stages of the exercise test, or 2) participant requested to stop the test, or 3) participant reached 85% of their age-predicted maximal HR. The TBRS submaximal exercise linear regression equation was then used to calculate the estimated maximal oxygen consumption (estimatedV□O_2max_).(25) The linear relationship between workload and HR (i.e. slope) was plotted and used to determine the estimated maximal watts (estimatedWatt_max_).(27)

#### Exercise Familiarization

After the TBRS submaximal exercise test, participants rested to allow HR and blood pressure to return to resting values. Individuals participated in the HIIE familiarization, switching between 1-minute high intensity and 1-minute low intensity intervals while maintaining a consistent step rate on the recumbent stepper. Based on previous protocols, intensity for the acute HIIE bout was calculated as 60-80% estimatedWatt_max_ for high intensity intervals and 10% estimatedWatt_max_ for low intensity intervals.(28–31) HR and RPE were measured at the end of each interval. Based on previous recumbent stepper HIIE protocols, a HR limit of 85% of age predicted maximum HR was used.(32) Participants also practiced breathing through their nose during the interval exercise. The familiarization session lasted approximately 10 minutes.

### Study Visit 2

#### Carotid Ultrasound

Upon arrival to the laboratory, participants underwent a carotid ultrasound scan. Participants rested quietly in a supine position for 20 minutes. We then conducted a bilateral Doppler ultrasound scan of the common, internal, and external carotid artery. Carotid 2D (GE Healthcare LOGIQ Ultrasound, Chicago, IL) images as well as systolic and diastolic velocity measures were sent to a collaborating physician (SXB) for carotid stenosis assessment.(22)

#### MCAv Recordings

After completing the carotid ultrasound scan, participants were seated on the recumbent stepper and rested quietly for 20 minutes while the following equipment was donned: 1) TCD headset and bilateral ultrasound probes to measure MCAv, 2) a 5-lead electrocardiogram (ECG; Cardiocard, Nasiff Associates, Central Square, New York) for HR, 3) a nasal cannula attached to a capnograph (BCI Capnocheck Sleep 9004 Smiths Medical, Dublin, Ohio) for expired end tidal carbon dioxide (P_ET_CO_2_) and respiratory rate (RR), 4) a beat-to-beat blood pressure cuff on the left middle finger (Finometer, Finapres Medical Systems, Amsterdam, the Netherlands) for mean arterial pressure (MAP), and 5) a brachial automated sphygmomanometer with a microphone on the right arm (Tango M2; Suntech, Morrisville, NC) to ensure an accurate calibration of the Finometer. The participant’s arms were placed on stable platforms at heart level. As in our previous work, the laboratory room was dimly lit and kept at a constant temperature of 22°C to 24°C.(5)

**Figure 1** details the experimental protocol. Following the BL rest recording, MCAv was continuously recorded during the 10-minute acute HIIE bout. The HIIE bout consisted of repetitive 1-minute high intensity intervals (60-80% estimatedWatt_max_) separated by 1-minute low intensity intervals (10% estimatedWatt_max_).(13, 29) The HIIE MCAv recording started with a 1-minute low intensity interval to prevent a Valsalva Maneuver, which can occur at the start of heavy resistance exercise, eliciting a large transient drop in MAP and MCAv.(33) After 10 minutes of HIIE, resistance was decreased to 15 watts and participants performed a 2-minute active cool-down. Immediately following the cool-down, participants stopped exercise and sat quietly on the recumbent stepper while we collected a 5-minute recording. The final recording lasted 5 minutes and was conducted 30 minutes after HIIE in a seated position.

**Figure 1.**
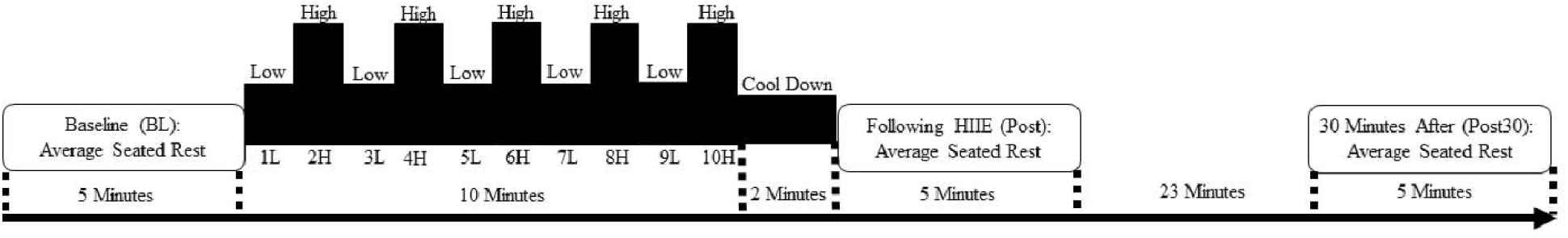
– Protocol Layout of MCAv recordings 1) before HIIE, 2) during HIIE, 3) following HIIE, and 4) 30 minutes after HIIE. L = Low intensity interval. H = High intensity interval.

**Figure 2 –.**
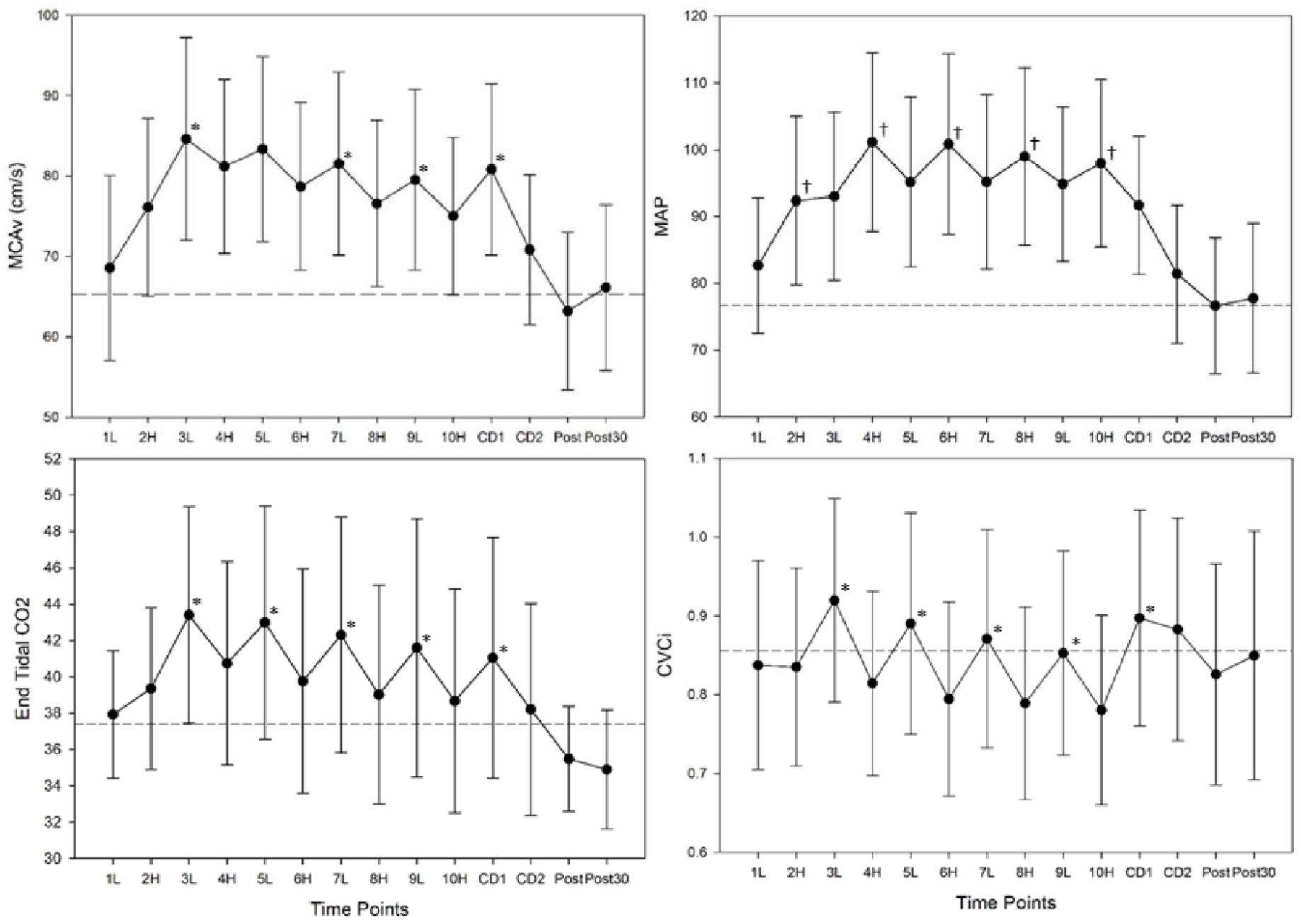
Response to high intensity interval exercise (HIIE, n = 14); Middle Cerebral Artery blood velocity (MCAv, cm·s^-1^), Mean Arterial Pressure (MAP, mm Hg), expired end tidal carbon dioxide (P_ET_CO_2_, mm Hg), and cerebrovascular conductance index (CVCi, MCAv/MAP). L = Low intensity interval. H = High intensity interval. Horizontal dashed line represents baseline value. *Indicates significant “rebounding” effect during low intensity compared to previous high intensity interval. ^†^ Indicates significant increase compared to previous low intensity interval.

Raw data were acquired through an analog-to-digital unit (NI-USB-6212, National Instruments) and custom-written software within MATLAB (v2014a, TheMathworks Inc, Natick, Massachusetts). Sampling rate was set at 500 Hz. Raw data were then time aligned and interpolated beat-to-beat using the R spike on EKG as a trigger. Each 5-minute rest recording was averaged separately at: 1)BL, 2) following HIIE, and 3) 30 minutes after HIIE. Each 1-minute interval during the 10-minute HIIE bout was averaged separately.

#### Data Analysis

Alpha was set a priori at 0.05. Normality was determined using a Shapiro-Wilk test. Consistent with previously published work, the left MCAv signal was used within the analysis to compare time points but if the left MCA signal was not obtainable, then right MCAv was used.(2, 15) To examine the conductance of peripheral blood pressure to the brain via MCAv we calculated the beat-to-beat cerebrovascular conductance index (CVCi = MCAv/MAP)(34). Participant characteristics were analyzed using a one-way ANOVA, Mann-Whitney U test for non-parametric variables, and Fisher’s exact test for categorical variables. Differences in the response to HIIE compared to BL were analyzed using a repeated measures ANOVA and post-hoc paired t-test with Bonferroni correction (α < 0.005). Relationships between the change in MCAv, MAP, and P_ET_CO_2_ were analyzed using a Spearman’s rank correlation coefficients. We used a repeated measures ANOVA with post-hoc paired t-test to detect differences following HIIE and 30-minutes after HIIE compared to BL. Differences in the MCAv response between sex were analyzed using a mixed design ANOVA and post-hoc independent t-test with Bonferroni correction.

## Results

### MCAv Response to HIIE

The average MCAv for each interval of HIIE is presented in **Table 2**. Odd integer intervals represent low intensity exercise while even integer intervals represent high intensity exercise, shown in **Figure 1**. Compared to BL, MCAv was significantly higher during intervals 2 through 10 of HIIE (p < 0.001). MCAv peaked at interval 3 and began to decrease throughout the remainder of the acute exercise bout, shown in **Table 2**. Following the high intensity intervals, we observed a “rebounding” response whereby MCAv increased during low intensity intervals, shown in **Figure 2**. Compared to BL, MCAv was also lower after HIIE (p = 0.03) but returned to BL values at 30-minutes after HIIE (p = 0.47), shown in **Table 3**.

**Table 2.** MCAv Response to HIIE.

|  | BL | 1 (L) | 2 (H) | 3 (L) | 4 (H) | 5 (L) | 6 (H) | 7 (L) | 8 (H) | 9 (L) | 10 (H) |
| --- | --- | --- | --- | --- | --- | --- | --- | --- | --- | --- | --- |
| **MCAv** | 65.1 ± | 68.6 ± | 76.1 ± | 84.6 ± | 81.2 ± | 83.4 ± | 78.7 ± | 81.5 ± | 76.6 ± | 79.5 ± | 75.0 ± |
| **(cm·s<sup>-1</sup>)** | 9.1 | 11.5 | 11.1* | 12.6* | 10.8* | 11.5* | 10.4* | 11.4* | 10.4* | 11.3* | 9.8* |
| **P<sub>ET</sub>CO<sub>2</sub>** | 37.3 ± | 37.9 ± | 39.3 ± | 43.4 ± | 40.7 ± | 43.0 ± | 39.8 ± | 42.3 ± | 39.0 ± | 41.6 ± | 38.7 ± |
| **(mmHg)** | 2.7 | 3.5 | 4.4 | 6.0* | 5.6 | 6.4* | 6.2 | 6.5* | 6.0 | 7.1 | 6.2 |
| **RR** | 14.0 ± | 21.2 ± | 23.3 ± | 20.2 ± | 23.9 ± | 20.8 ± | 24.5 ± | 21.5 ± | 24.9 ± | 22.5 ± | 25.5 ± |
| **(br·min<sup>-1</sup>)** | 3.2 | 3.0* | 3.5* | 4.9* | 4.6* | 4.4* | 4.6* | 4.4* | 4.7* | 4.5* | 4.7* |
| **MAP** | 76.7 ± | 82.7 ± | 92.4 ± | 93.0 ± | 101.1 ± | 95.2 ± | 100.9 ± | 95.2 ± | 99.0 ± | 94.8 ± | 98.0 ± |
| **(mmHg)** | 11.3 | 10.2* | 12.6* | 12.6* | 13.4* | 12.7* | 13.5* | 13.1* | 13.3* | 11.6* | 12.5* |
| **CVCi** | 0.87 ± | 0.84 ± | 0.84 ± | 0.92 ± | 0.81 ± | 0.89 ± | 0.79 ± | 0.87 ± | 0.79 ± | 0.85 ± | 0.78 ± |
|  | 0.2 | 0.1 | 0.1 | 0.1 | 0.1 | 0.1 | 0.1 | 0.1 | 0.1 | 0.1 | 0.1 |
| **PeakHR** | - | 47% ± | 65% ± | 54% ± | 69% ± | 57% ± | 72% ± | 60% | 74% ± | 62% ± | 75% ± |
| **(%max)** |  | 8% | 6% | 8% | 7% | 9% | 7% | ± 9% | 7% | 9% | 7% |

Mean ± standard deviations. Due to repeated measures with multiple comparisons, $\alpha$ was set at 0.005.
\* = Significantly different than Baseline. MCAv = Middle Cerebral Artery Blood Velocity. P<sub>ET</sub>CO<sub>2</sub> = End Tidal Carbon Dioxide. RR = Respiratory Rate. br·min<sup>-1</sup> = breaths per minute. MAP = Mean Arterial Pressure. CVCi = Cerebrovascular Conductance Index. PeakHR = Peak Heart Rate. %max = Percent Age Predicted Maximum Heart Rate.

**Table 3.** MCAv Recovery Response to HIIE.

|  | BL | Post | Post 30 |
| --- | --- | --- | --- |
| **MCAv ($\text{cm}\cdot\text{s}^{-1}$)** | $65.1 \pm 9.1$ | $63.2 \pm 9.8^*$ | $66.1 \pm 10.3$ |
| **P<sub>ET</sub>CO<sub>2</sub> (mmHg)** | $37.3 \pm 2.7$ | $35.5 \pm 2.9^*$ | $34.9 \pm 3.3^*$ |
| **RR ($\text{br}\cdot\text{min}^{-1}$)** | $14.0 \pm 3.2$ | $16.1 \pm 3.5^*$ | $14.9 \pm 3.1$ |
| **MAP (mmHg)** | $76.7 \pm 11.3$ | $76.6 \pm 10.2$ | $77.7 \pm 11.2$ |
| **CVCi** | $0.87 \pm 0.2$ | $0.84 \pm 0.1$ | $0.86 \pm 0.2$ |
| **HR (bpm)** | $72 \pm 11$ | $96 \pm 17^*$ | $83 \pm 12^*$ |

Mean ± Standard Deviations. A priori $\alpha = 0.05$ . \* = Significantly different than
Baseline. MCAv = Middle Cerebral Artery Blood Velocity. P<sub>ET</sub>CO<sub>2</sub> = End Tidal
Carbon Dioxide. RR = Respiratory Rate. $\text{br}\cdot\text{min}^{-1}$ = breaths per minute. MAP = Mean
Arterial Pressure. CVCi = Cerebrovascular Conductance Index. HR = Heart Rate

### MAP, HR, P_ET_CO_2_ and CVCi Response to HIIE

During HIIE, MAP increased significantly from BL values (p < 0.002), shown in **Table 2**. MAP also significantly increased during the high intensity intervals compared to low intensity intervals (p < 0.005), shown in **Figure 2**. The change in MAP was significantly related to the change in MCAv between intervals 2 and 3 (r = 0.82, p < 0.001), intervals 3 and 4 (r = 0.62, p = 0.02), and intervals 8 and 9 (r = 0.54, p = 0.045). MAP returned to resting values and was not significantly different from BL, following HIIE (p = 0.97) or 30-minutes after HIIE (p = 0.75), shown in **Table 3**.

Compared to BL, P_ET_CO_2_ increased significantly during low intensity intervals 3, 5 and 7 (p <0.003), shown in **Table 2**. Compared to BL, RR increased during HIIE and was increased during high intensity compared to low intensity intervals (p ≤ 0.003). Compared to interval 3 (the peak MCAv interval), RR increased during high intensity intervals (p <0.001), which is consistent with the pattern of hyperventilation induced decreases in MCAv shown in **Figure 2**.

Due to increased RR (hyperventilation) during the high intensity intervals, P_ET_CO_2_ was not significantly different from BL during high intensity. The change in P_ET_CO_2_ was significantly related to the change in MCAv between intervals 1 and 2 (r = 0.60, p = 0.03), intervals 6 and 7 (r = 0.69, p = 0.01), intervals 7 and 8 (r = 0.81, p <0.001), intervals 8 and 9 (r = 0.53, p = 0.049), and intervals 9 and 10 (r = 0.65, p = 0.01). During recovery, P_ET_CO_2_ was significantly lower than BL following HIIE (p = 0.002) and 30-minutes after HIIE (p = 0.02). RR was lower than BL following HIIE (p = 0.02) but returned to BL levels at 30-minutes after HIIE (p = 0.16).

CVCi did not significantly change from BL during the acute HIIE bout, following HIIE or 30-minutes after HIIE. Following interval 3 (the peak MCAv interval), CVCi decreased during high intensity intervals and increased with low intensity intervals (p < 0.001), shown in **Figure 3**. The slope from interval onset to peak CVCi was calculated during interval 1, 2, 3, 5, 7, and 9. With CVCi decreasing during intervals 4, 6, 8, and 10, the slope from interval onset to nadir CVCi value was calculated. The slopes of the CVCi response were different between high and low intensity intervals following interval 3 (p < 0.001). The first high intensity interval, or interval 2, was faster to peak CVCi (18 ± 15 seconds) compared to the other high intensity intervals (p ≤ 0.001). The time to peak or nadir CVCi was also significantly faster during low intensity intervals compared to high intensity intervals such as: interval 5 and 6 (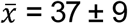 seconds versus 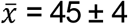 seconds, p = 0.02), and interval 7 and 8 (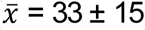 seconds versus 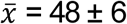 seconds, p = 0.02).

**Figure 3–.**
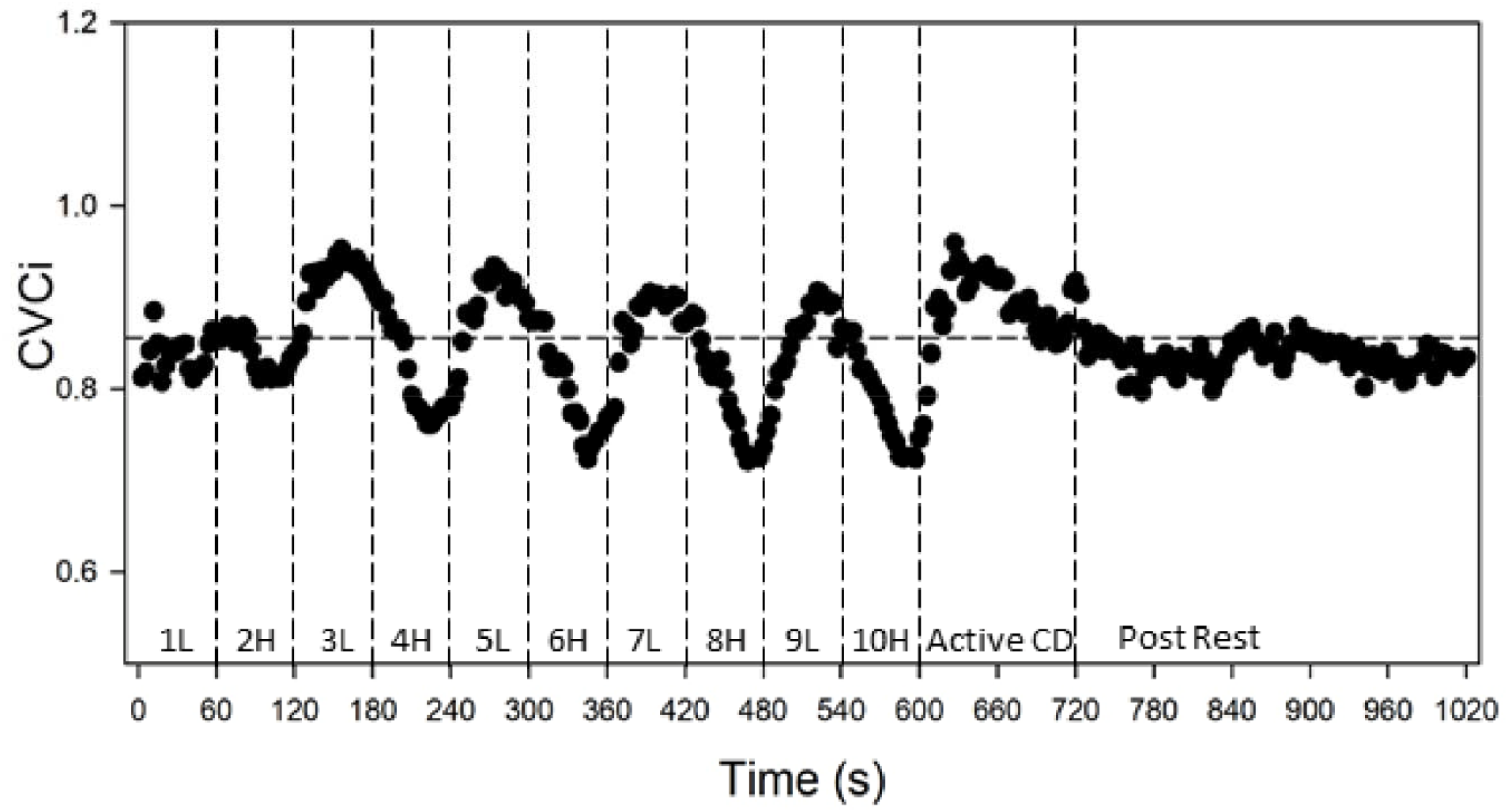
Cerebrovascular conductance index (CVCi, MCAv/MAP) with each heartbeat. L = Low intensity interval H = High intensity interval. CD = Cool Down. Horizontal dashed line represents baseline value. High intensity interval exercise (HIIE) occurred during 0 – 600 seconds (10 minutes). Active cool down occurred between 600-720 seconds (2 minutes). Passive recovery following HIIE occurred between 720 – 1020 seconds (5 minutes).

### Sex Differences in MCAv Response to HIIE

The MCAv response to HIIE differed between men and women, shown in **Table 4**. Although resting BL MCAv was significantly higher in women compared to men (p = 0.01), the percent change in MCAv was greater in men, shown in **Figure 4**. Therefore, during intervals 4 through 10, MCAv was no longer significantly different between men and women. However, following HIIE and 30-minutes after HIIE, MCAv returned to being higher in women compared to men (p = 0.01). In women, the change in MCAv was related to the change in P_ET_CO_2_ between intervals 4 and 5 (r = 0.81, p = 0.03), intervals 6 and 7 (r = 0.90, p = 0.01), intervals 7 and 8 (r = 0.87, p = 0.01), and intervals 9 and 10 (r = 0.87, p = 0.01). In men, the change in MCAv was related to the change in P_ET_CO_2_ between intervals 6 and 7 (r = 0.75, p = 0.05) and intervals 7 and 8 (r = 0.98, p < 0.001).

**Table 4.**
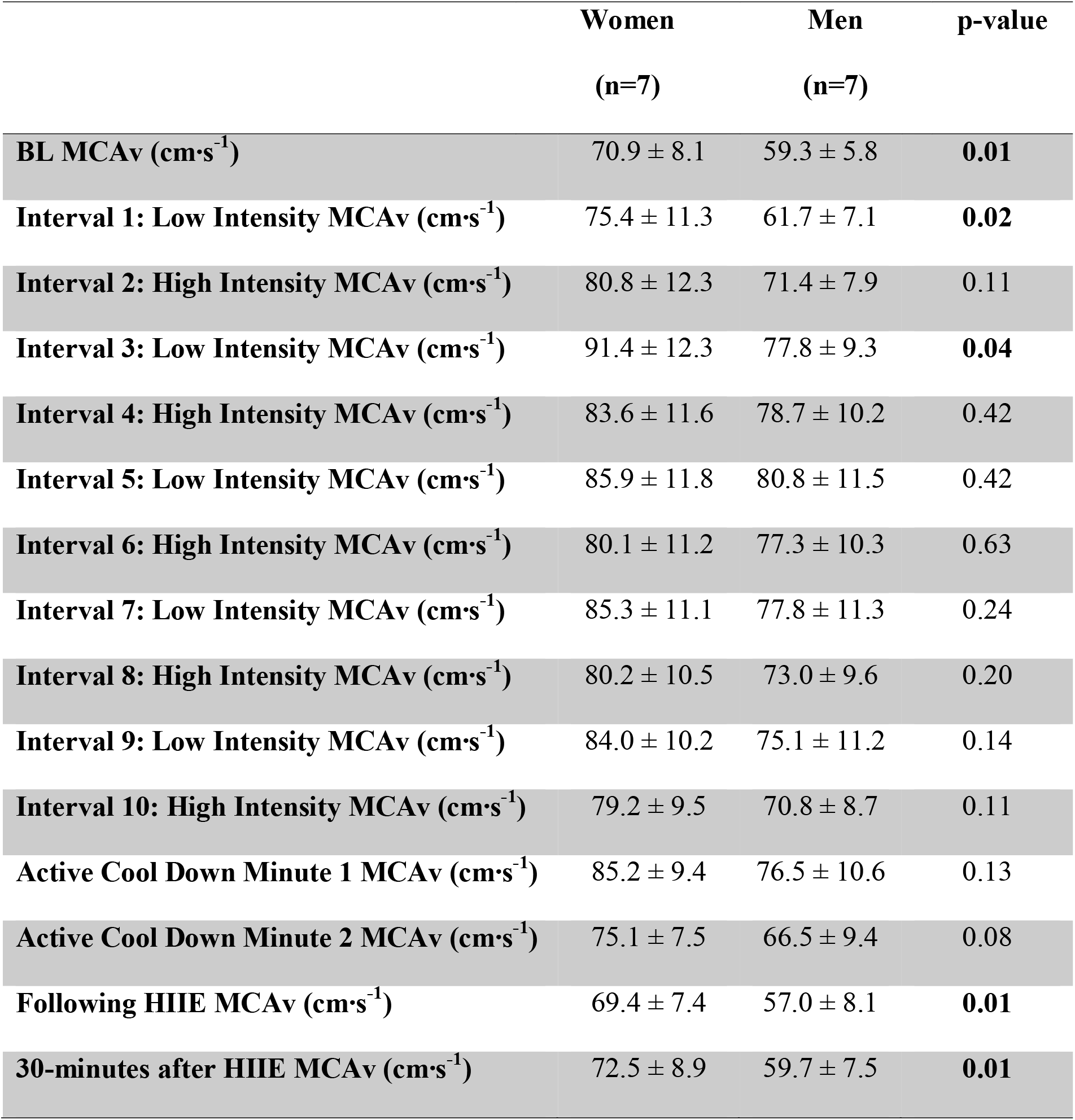
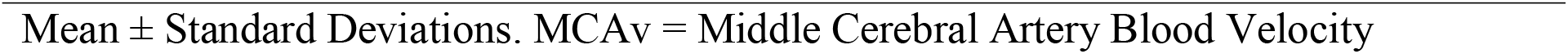
Absolute MCAv Response Between Sex.

**Figure 4–.**
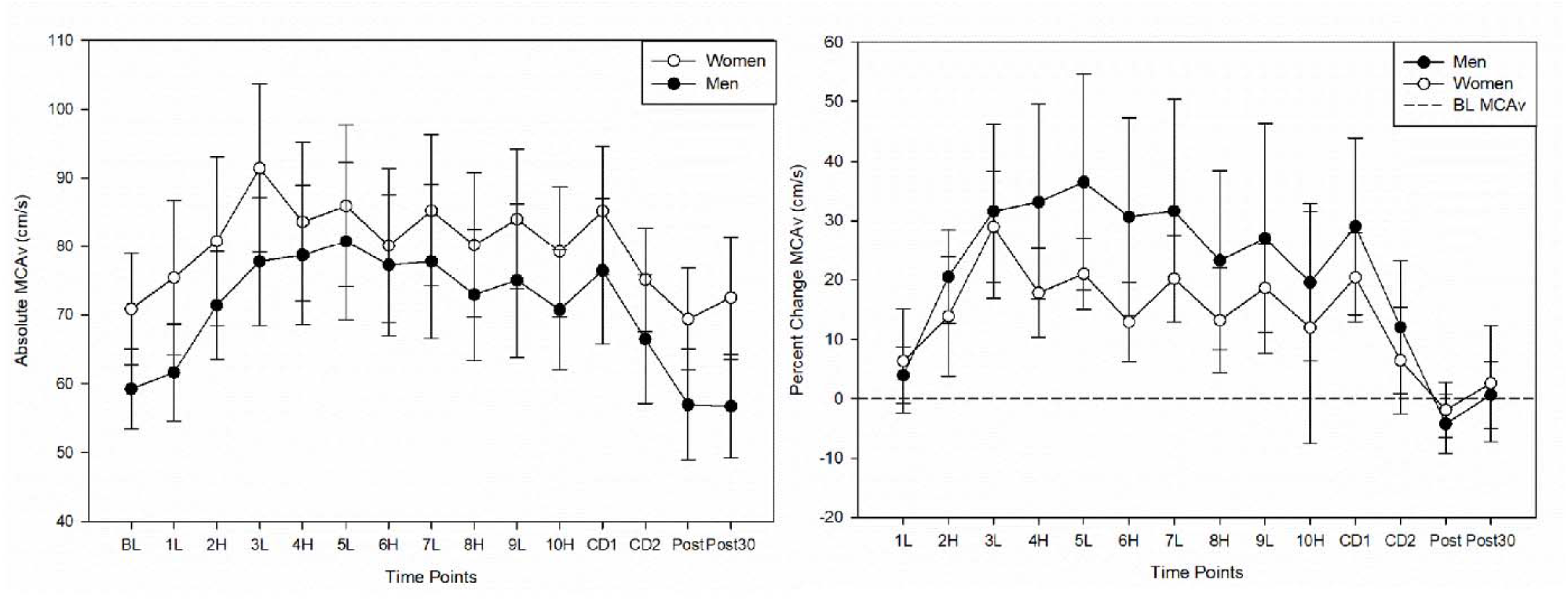
Absolute MCAv response to high intensity interval exercise (left) compared to the relative percent change MCAv response (left). Horizontal dashed line represents baseline value.

## Discussion

Short interval HIIE is widely utilized to improve peripheral vascular health(35, 36) despite minimal research into its effects on the cerebrovascular system.(10) This study provides novel and crucial insight into the MCAv response to rapid and repetitive short interval HIIE in healthy young adults. The primary findings of the study are: 1) MCAv did not decrease below resting values during HIIE, 2) MCAv peaked within 3 minutes of HIIE and began to decrease with high intensity, 3) MCAv rebounded during the low intensity intervals, 4) MCAv decreased below resting values for 5 minutes after HIIE, and 5) resting MCAv was higher in women compared to men, but men had a greater change in MCAv during HIIE.

The primary objective of this study was to characterize the MCAv response during an acute bout of 1-minute interval HIIE in healthy adults. Based on previously published work,(11) we hypothesized that MCAv would decrease below BL MCAv during an acute 10-minute HIIE bout. However, our primary hypothesis was not supported by our findings. While MCAv did not decrease below resting MCAv throughout the HIIE bout, MCAv did start to decrease after 3 minutes of HIIE. Initially, MCAv increased with the onset of HIIE and was moderately correlated with the change in MAP. However, MCAv began to decrease after 3 minutes of HIIE and was moderately correlated with the change in P_ET_CO_2_. Therefore, the initial rise in MCAv could be partially explained by peripheral blood pressure while the latter decrease in MCAv was driven by hyperventilation induced arteriole vasoconstriction.

Prior research has demonstrated a decrease in MCAv below BL resting MCAv during 1-minute interval HIIE in young children.(11) In contrast, we did not find a reduction in MCAv below BL MCAv, which could be due to the age of the participants (children versus adults), the high intensity workload (90% versus 70% estimated watt maximum), and the exercise modality (cycle versus recumbent stepper). Our results are supported by previous research performing short sprint interval training in healthy adults that also found no reduction in MCAv below BL resting MCAv.(12) However, the prior study performed 30-second sprint intervals at 200% watt maximum with longer 4.5-minute recovery intervals. Therefore, our study contributes to the existing literature showing no reduction in MCAv below BL during an acute 10-minute bout of short interval HIIE with alternating 1-minute high intensity and low intensity intervals.

While MCAv started to decrease after 3 minutes of HIIE during the high intensity intervals, MCAv increased or “rebounded” during the low intensity intervals. Previous studies measuring MCAv during HIIE have also reported this “rebounding” effect.(11, 12, 34) MCAv “rebounding” during light intensity intervals could be partially explained by P_ET_CO_2_ as they were moderately correlated. MCAv also closely followed the pattern of P_ET_CO_2,_ with both measures decreasing during high intensity (hyperventilation) and increasing during recovery. We have also shown increased CVCi during the recovery intervals, showing a potential contribution of MAP to the “rebounding effect”. It has been hypothesized that the “rebounding effect” could due to the increase in CVCi, or conductance of MAP to the brain, due to an attenuation of cerebral autoregulation (ability of the brain to maintain homeostasis during blood pressure fluctuations) with an acute bout of high intensity exercise.(12, 18, 34) Others have hypothesized that increased shear stress on the cerebrovascular arterial walls with high intensity exercise could create vasodilatory effects.(12, 37) However, future research measuring dynamic cerebrovascular autoregulation and the cerebrovascular flow mediated dilatory response to short interval HIIE requires further exploration.

Our secondary hypotheses were that MCAv would be lower than BL MCAv during passive recovery 1) following HIIE and 2) 30-minutes after HIIE. Our hypotheses were partially supported by our findings. MCAv was lower during the 5-minute rest following HIIE but not at 30-minutes after HIIE. Our findings are supported by previous research also reporting a reduction in MCAv below BL following HIIE.(11) The reduction in MCAv following exercise may be a unique characteristic to HIIE, as a previous study performing 20 minutes of moderate intensity continuous exercise reported no reduction in MCAv after exercise in healthy adults.(38)

The reduction in MCAv cannot be fully explained by P_ET_CO_2_ considering MCAv recovers to BL resting values at 30-minutes after HIIE while P_ET_CO_2_ remains significantly decreased.(11) This could be due to an attenuation of cerebrovascular reactivity to carbon dioxide following an acute bout of HIIE.(10, 39)

### CVCi Response

We are the first to characterize the dynamic CVCi response during HIIE. We report a quick increase in conductance during the first high intensity interval with a subsequently longer time for the remaining high intensity intervals. Similar to Labrecque et al. examining the time to peak MCAv during a 30-second high intensity sprint,(34) we characterized the time to peak (or nadir) CVCi during each 1-minute high intensity and low intensity intervals. We found that the time to peak CVCi was fastest during the first high intensity interval compared to the remaining high intensity intervals. We hypothesize that the longer time to peak/nadir CVCi during high intensity intervals could be a regulatory response attenuating quick fluctuations in CVCi with increased MAP. The slope of the CVCi response was negative during the latter high intensity intervals, due to hyperventilation induced decreases in MCAv. However, CVCi increased or “rebounded” during the low intensity intervals. While prior work has shown a decreased regulatory response following HIIE,(18) further research is needed to determine whether the increase in CVCi or “rebounding effect” is due to an attenuation of the regulatory response during HIIE.

### Sex Differences

Our previous work identified women as having a higher BL resting MCAv compared to men but no significant difference in the MCAv response to moderate intensity continuous exercise.(14, 15) While previous work has been published on the MCAv response to interval exercise, to our knowledge, no one has studied sex differences during short interval HIIE and recovery.(12, 13) Although women had a higher BL MCAv at rest, MCAv was not significantly different during exercise due to a larger MCAv response in men. As observed in **Table 1**, the men worked at a significantly greater absolute workload than women, which we propose influenced the greater MCAv response. The MCAv differences between sex could also be due to young men having a higher resting pulsatility index, as lower MCAv has been hypothesized to be a protective mechanism for the downstream microvasculature.(14, 40) Another potential explanation for the lack of significance during HIIE could be due to women having a greater reactivity to carbon dioxide causing decreases in MCAv, which is supported by strong correlations between the change in MCAv and change in P_ET_CO_2_ in women compared to men.(41) Although we standardized the visits to be within days 1-7 of the follicular phase, we did not directly measure the effects of hormones on MCAv in both women and men in this study.

### Limitations

There are limitations for the current study. First, the middle cerebral artery (MCA) diameter could not be directly measured. There is controversy whether MCA diameter is vasoactive with high intensity exercise induced hyperventilation.(42, 43) One study states that MCAv may underestimate changes in cerebral blood flow during changes in P_ET_CO_2_,(44) while another study reports no change in MCA diameter measured through magnetic resonance imaging with a P_ET_CO_2_ change of 7.5mm Hg during hyperventilation.(42) However, due to the difficulty of temporally assessing MCA diameter during high intensity exercise, we assumed the MCA diameter remains constant. Second, HIIE was dosed based on an estimated maximal intensity rather than directly measuring the anerobic threshold. However, based on the P_ET_CO_2_ response, we believe the HIIE was accurately prescribed. Third, three individuals were also outside the window of 2-14 days between study visits. However, these individuals confirmed no changes to their health or physical activity routine between study visits 1 and 2. Therefore, in the context of this study design in healthy adults, the results were not likely skewed. Lastly, P_ET_CO_2_ was measured via nasal cannula, and breathing strictly through the nose during HIIE could become increasingly difficult. However, a study team member monitored nasal breathing throughout the entirety of the MCAv recordings and acute HIIE bout to ensure adherence to the protocol and appropriate P_ET_CO_2_ values.

### Future Considerations

Prior literature reviews suggest HIIE results in rapid changes (oscillations) in blood pressure that could challenge the cerebrovascular system, particularly those with cerebrovascular impairment.(45) Despite these proposed considerations, high intensity gait training and interval exercise are recommended to improve walking for those with neurological disease.(46) Further, clinical practice guidelines are beginning to include HIIE with short intervals of 30-seconds or 1-minute to improve walking outcomes after stroke.(28, 46, 47) The logical next step in this line of research will be to focus on clinical population such as stroke.

## Conclusion

During an acute 10-minute bout of HIIE with short intervals, MCAv does not decrease below BL resting MCAv in healthy young adults. However, MCAv falls below BL resting MCAv following HIIE but returns to BL at 30-minutes after HIIE. Further exploration into the regulatory response and physiological mechanisms contributing to a reduction in MCAv during passive recovery after HIIE is needed. This study contributes novel characterizations of the MCAv response to short interval HIIE in healthy adult controls, including changes in CVCi and sex differences. We have laid the foundation for studying the cerebrovascular response to short interval HIIE.(46)

## Supporting information

Strobe Checklist

COI Disclosure

COI Disclosure

COI Disclosure

COI Disclosure

COI Disclosure

COI Disclosure

COI Disclosure

## Data Availability

Data is available upon request to authors.

## Funding and Acknowledgements

We would like to thank Katherine Nguyen and Jake Buchanan for their contributions to data collection. AW was supported by the Eunice Kennedy Shriver National Institute of Child Health & Human Development of the National Institutes of Health (T32HD057850). The content is solely the responsibility of the authors and does not necessarily represent the official views of the National Institutes of Health. REDCap at University of Kansas Medical Center is supported by Clinical and Translational Science Awards (CTSA) Award # ULTR000001 from National Center for Research Resources (NCRR). The Georgia Holland Research in Exercise and Cardiovascular Health (REACH) laboratory space was supported by the Georgia Holland Endowment Fund.

## Conflicts of Interest

The author(s) report no conflict of interest. The results of this study have been presented clearly, honestly, and without fabrication, falsification, or inappropriate data manipulation.

## Notes

### Clinical Trial

NCT04673994

### Author Declarations

The study and all experimental procedures were approved by the University of Kansas Medical Center Human Subjects Committee.

## References

1. Smith KJ, Ainslie PN. Regulation of cerebral blood flow and metabolism during exercise. Exp Physiol. 2017;102(11):1356–71. Epub 2017/08/09. doi: 10.1113/EP086249. PubMed PMID: 28786150.

2. Witte E, Liu Y, Ward JL, Kempf KS, Whitaker A, Vidoni ED, et al. Exercise intensity and middle cerebral artery dynamics in humans. Respir Physiol Neurobiol. 2019;262:32–9. Epub 2019/02/03. doi: 10.1016/j.resp.2019.01.013. PubMed PMID: 30710650; PubMed Central PMCID: PMCPMC6393201.

3. Thomas SN, Schroeder T, Secher NH, Mitchell JH. Cerebral blood flow during submaximal and maximal dynamic exercise in humans. J Appl Physiol (1985). 1989;67(2):744–8. Epub 1989/08/01. doi: 10.1152/jappl.1989.67.2.744. PubMed PMID: 2507500.

4. Hellstrom G, Wahlgren NG. Physical exercise increases middle cerebral artery blood flow velocity. Neurosurg Rev. 1993;16(2):151–6. Epub 1993/01/01. doi: 10.1007/BF00258249. PubMed PMID: 8345909.

5. Billinger SA, Craig JC, Kwapiszeski SJ, Sisante JV, Vidoni ED, Maletsky R, et al. Dynamics of middle cerebral artery blood flow velocity during moderate-intensity exercise. J Appl Physiol (1985). 2017;122(5):1125–33. Epub 2017/03/11. doi: 10.1152/japplphysiol.00995.2016. PubMed PMID: 28280106; PubMed Central PMCID: PMCPMC5451537.

6. Olson TP TJ, Dengel DR. Relationship between ventilatory threshold and cerebral blood flow during maximal exercise in humans. Open Sports Med J. 2009;3(9):13. doi: 10.2174/1874387000903010009.

7. Jorgensen LG, Perko G, Secher NH. Regional cerebral artery mean flow velocity and blood flow during dynamic exercise in humans. J Appl Physiol (1985). 1992;73(5):1825–30. Epub 1992/11/01. doi: 10.1152/jappl.1992.73.5.1825. PubMed PMID: 1474058.

8. Fitts RH. Cellular mechanisms of muscle fatigue. Physiol Rev. 1994;74(1):49–94. Epub 1994/01/01. doi: 10.1152/physrev.1994.74.1.49. PubMed PMID: 8295935.

9. Ainslie PN, Duffin J. Integration of cerebrovascular CO2 reactivity and chemoreflex control of breathing: mechanisms of regulation, measurement, and interpretation. Am J Physiol Regul Integr Comp Physiol. 2009;296(5):R1473–95. Epub 2009/02/13. doi: 10.1152/ajpregu.91008.2008. PubMed PMID: 19211719.

10. Whitaker AA, Alwatban M, Freemyer A, Perales-Puchalt J, Billinger SA. Effects of high intensity interval exercise on cerebrovascular function: A systematic review. PLoS One. 2020;15(10):e0241248. Epub 2020/10/30. doi: 10.1371/journal.pone.0241248. PubMed PMID: 33119691.

11. Tallon CM, Simair RG, Koziol AV, Ainslie PN, McManus AM. Intracranial Vascular Responses to High-Intensity Interval Exercise and Moderate-Intensity Steady-State Exercise in Children. Pediatr Exerc Sci. 2019;31(3):290–5. Epub 2019/03/06. doi: 10.1123/pes.2018-0234. PubMed PMID: 30832540.

12. Weaver SR, Skinner BD, Furlong R, Lucas RAI, Cable NT, Rendeiro C, et al. Cerebral Hemodynamic and Neurotrophic Factor Responses Are Dependent on the Type of Exercise. Front Physiol. 2020;11:609935. Epub 2021/02/09. doi: 10.3389/fphys.2020.609935. PubMed PMID: 33551835; PubMed Central PMCID: PMCPMC7859714.

13. Klein T, Bailey TG, Abeln V, Schneider S, Askew CD. Cerebral Blood Flow during Interval and Continuous Exercise in Young and Old Men. Med Sci Sports Exerc. 2019;51(7):1523–31. Epub 2019/02/16. doi: 10.1249/MSS.0000000000001924. PubMed PMID: 30768552.

14. Alwatban MR, Aaron SE, Kaufman CS, Barnes JN, Brassard P, Ward JL, et al. Effects of Age and Sex on Middle Cerebral Artery Blood Velocity and Flow Pulsatility Index Across the Adult Lifespan. J Appl Physiol (1985). 2021. Epub 2021/03/12. doi: 10.1152/japplphysiol.00926.2020. PubMed PMID: 33703940.

15. Ward JL, Craig JC, Liu Y, Vidoni ED, Maletsky R, Poole DC, et al. Effect of healthy aging and sex on middle cerebral artery blood velocity dynamics during moderate-intensity exercise. Am J Physiol Heart Circ Physiol. 2018;315(3):H492–H501. Epub 2018/05/19. doi: 10.1152/ajpheart.00129.2018. PubMed PMID: 29775407; PubMed Central PMCID: PMCPMC6172645.

16. Thompson PD, Arena R, Riebe D, Pescatello LS, American College of Sports M. ACSM’s new preparticipation health screening recommendations from ACSM’s guidelines for exercise testing and prescription, ninth edition. Curr Sports Med Rep. 2013;12(4):215–7. Epub 2013/07/16. doi: 10.1249/JSR.0b013e31829a68cf. PubMed PMID: 23851406.

17. Perod AL, Roberts AE, McKinney WM. Caffeine can affect velocity in the middle cerebral artery during hyperventilation, hypoventilation, and thinking: a transcranial Doppler study. J Neuroimaging. 2000;10(1):33–8. Epub 2000/02/10. doi: 10.1111/jon200010133. PubMed PMID: 10666980.

18. Burma JS, Copeland P, Macaulay A, Khatra O, Wright AD, Smirl JD. Dynamic cerebral autoregulation across the cardiac cycle during 8 hr of recovery from acute exercise. Physiol Rep. 2020;8(5):e14367. Epub 2020/03/13. doi: 10.14814/phy2.14367. PubMed PMID: 32163235; PubMed Central PMCID: PMCPMC7066871.

19. Mathew RJ, Wilson WH. Regional cerebral blood flow changes associated with ethanol intoxication. Stroke. 1986;17(6):1156–9. Epub 1986/11/01. doi: 10.1161/01.str.17.6.1156. PubMed PMID: 3810714.

20. Krejza J, Mariak Z, Huba M, Wolczynski S, Lewko J. Effect of endogenous estrogen on blood flow through carotid arteries. Stroke. 2001;32(1):30–6. Epub 2001/01/04. doi: 10.1161/01.str.32.1.30. PubMed PMID: 11136910.

21. Conroy DA, Spielman AJ, Scott RQ. Daily rhythm of cerebral blood flow velocity. J Circadian Rhythms. 2005;3(1):3. Epub 2005/03/12. doi: 10.1186/1740-3391-3-3. PubMed PMID: 15760472; PubMed Central PMCID: PMCPMC555580.

22. Grant EG, Benson CB, Moneta GL, Alexandrov AV, Baker JD, Bluth EI, et al. Carotid artery stenosis: gray-scale and Doppler US diagnosis--Society of Radiologists in Ultrasound Consensus Conference. Radiology. 2003;229(2):340–6. Epub 2003/09/23. doi: 10.1148/radiol.2292030516. PubMed PMID: 14500855.

23. Whitfield GP, Pettee Gabriel KK, Rahbar MH, Kohl HW, 3rd. Application of the American Heart Association/American College of Sports Medicine Adult Preparticipation Screening Checklist to a nationally representative sample of US adults aged >=40 years from the National Health and Nutrition Examination Survey 2001 to 2004. Circulation. 2014;129(10):1113–20. Epub 2014/01/15. doi: 10.1161/CIRCULATIONAHA.113.004160. PubMed PMID: 24421370; PubMed Central PMCID: PMCPMC4094111.

24. Alexandrov AV, Sloan MA, Wong LK, Douville C, Razumovsky AY, Koroshetz WJ, et al. Practice standards for transcranial Doppler ultrasound: part I--test performance. J Neuroimaging. 2007;17(1):11–8. Epub 2007/01/24. doi: 10.1111/j.1552-6569.2006.00088.x. PubMed PMID: 17238867.

25. Billinger SA, E Vans, McClain M, Lentz AA, Good MB. Recumbent stepper submaximal exercise test to predict peak oxygen uptake. Med Sci Sports Exerc. 2012;44(8):1539–44. Epub 2012/03/03. doi: 10.1249/MSS.0b013e31824f5be4. PubMed PMID: 22382170; PubMed Central PMCID: PMCPMC3388171.

26. Wilson DR, Mattlage AE, Seier NM, Todd JD, Price BG, Kwapiszeski SJ, et al. Recumbent Stepper Submaximal Test response is reliable in adults with and without stroke. PLoS One. 2017;12(2):e0172294. Epub 2017/02/17. doi: 10.1371/journal.pone.0172294. PubMed PMID: 28207854; PubMed Central PMCID: PMCPMC5312932.

27. YMCA. Y’s Way to Physical Fitness 3rd Edition ed1989.

28. Boyne P, Dunning K, Carl D, Gerson M, Khoury J, Rockwell B, et al. High-Intensity Interval Training and Moderate-Intensity Continuous Training in Ambulatory Chronic Stroke: Feasibility Study. Phys Ther. 2016;96(10):1533–44. Epub 2016/04/23. doi: 10.2522/ptj.20150277. PubMed PMID: 27103222; PubMed Central PMCID: PMCPMC5046191.

29. Currie KD, Dubberley JB, McKelvie RS, MacDonald MJ. Low-volume, high-intensity interval training in patients with CAD. Med Sci Sports Exerc. 2013;45(8):1436–42. Epub 2013/03/09. doi: 10.1249/MSS.0b013e31828bbbd4. PubMed PMID: 23470301.

30. Nepveu JF, Thiel A, Tang A, Fung J, Lundbye-Jensen J, Boyd LA, et al. A Single Bout of High-Intensity Interval Training Improves Motor Skill Retention in Individuals With Stroke. Neurorehabil Neural Repair. 2017;31(8):726–35. Epub 2017/07/12. doi: 10.1177/1545968317718269. PubMed PMID: 28691645.

31. Crozier J, Roig M, Eng JJ, MacKay-Lyons M, Fung J, Ploughman M, et al. High-Intensity Interval Training After Stroke: An Opportunity to Promote Functional Recovery, Cardiovascular Health, and Neuroplasticity. Neurorehabil Neural Repair. 2018;32(6-7):543–56. Epub 2018/04/21. doi: 10.1177/1545968318766663. PubMed PMID: 29676956.

32. Boyne P MC, Carl D, Wilkerson J, Dunning K. Feasibility, Intensity and Safety of Recumbent Stepper High-intensity Interval Training in Chronic Stroke. Arch Phys Med Rehabil. 2015;96(12).

33. Perry BG, Cotter JD, Mejuto G, Mundel T, Lucas SJ. Cerebral hemodynamics during graded Valsalva maneuvers. Front Physiol. 2014;5:349. Epub 2014/10/14. doi: 10.3389/fphys.2014.00349. PubMed PMID: 25309449; PubMed Central PMCID: PMCPMC4163977.

34. Labrecque L, Drapeau A, Rahimaly K, Imhoff S, Billaut F, Brassard P. Comparable blood velocity changes in middle and posterior cerebral arteries during and following acute high-intensity exercise in young fit women. Physiol Rep. 2020;8(9):e14430. Epub 2020/04/29. doi: 10.14814/phy2.14430. PubMed PMID: 32342622; PubMed Central PMCID: PMCPMC7186567.

35. Weston KS, Wisloff U, Coombes JS. High-intensity interval training in patients with lifestyle-induced cardiometabolic disease: a systematic review and meta-analysis. Br J Sports Med. 2014;48(16):1227–34. Epub 2013/10/23. doi: 10.1136/bjsports-2013-092576. PubMed PMID: 24144531.

36. Ramirez-Velez R, Hernandez-Quinones PA, Tordecilla-Sanders A, Alvarez C, Ramirez-Campillo R, Izquierdo M, et al. Effectiveness of HIIT compared to moderate continuous training in improving vascular parameters in inactive adults. Lipids Health Dis. 2019;18(1):42. Epub 2019/02/06. doi: 10.1186/s12944-019-0981-z. PubMed PMID: 30717757; PubMed Central PMCID: PMCPMC6362599.

37. Carr J, Hoiland RL, Caldwell HG, Coombs GB, Howe CA, Tremblay JC, et al. Internal carotid and brachial artery shear-dependent vasodilator function in young healthy humans. J Physiol. 2020;598(23):5333–50. Epub 2020/09/10. doi: 10.1113/JP280369. PubMed PMID: 32901919.

38. Robertson AD, Atwi S, Kostoglou K, Verhoeff N, Oh PI, Mitsis GD, et al. Cerebrovascular Pulsatility During Rest and Exercise Reflects Hemodynamic Impairment in Stroke and Cerebral Small Vessel Disease. Ultrasound Med Biol. 2019;45(12):3116–27. Epub 2019/10/02. doi: 10.1016/j.ultrasmedbio.2019.08.019. PubMed PMID: 31570171.

39. Burma JS, Macaulay A, Copeland P, Khatra O, Bouliane KJ, Smirl JD. Comparison of cerebrovascular reactivity recovery following high-intensity interval training and moderate-intensity continuous training. Physiol Rep. 2020;8(11):e14467. Epub 2020/06/09. doi: 10.14814/phy2.14467. PubMed PMID: 32506845; PubMed Central PMCID: PMCPMC7276190.

40. Alwatban MR, Liu Y, Perdomo SJ, Ward JL, Vidoni ED, Burns JM, et al. TCD Cerebral Hemodynamic Changes during Moderate-Intensity Exercise in Older Adults. J Neuroimaging. 2020;30(1):76–81. Epub 2019/11/22. doi: 10.1111/jon.12675. PubMed PMID: 31750593; PubMed Central PMCID: PMCPMC6954976.

41. Kastrup A, Thomas C, Hartmann C, Schabet M. Sex dependency of cerebrovascular CO2 reactivity in normal subjects. Stroke. 1997;28(12):2353–6. Epub 1997/12/31. doi: 10.1161/01.str.28.12.2353. PubMed PMID: 9412613.

42. Valdueza JM, Balzer JO, Villringer A, Vogl TJ, Kutter R, Einhaupl KM. Changes in blood flow velocity and diameter of the middle cerebral artery during hyperventilation: assessment with MR and transcranial Doppler sonography. AJNR Am J Neuroradiol. 1997;18(10):1929–34. Epub 1997/12/24. PubMed PMID: 9403456.

43. Hoiland RL, Ainslie PN. CrossTalk proposal: The middle cerebral artery diameter does change during alterations in arterial blood gases and blood pressure. J Physiol. 2016;594(15):4073–5. Epub 2016/03/25. doi: 10.1113/JP271981. PubMed PMID: 27010010; PubMed Central PMCID: PMCPMC4806217.

44. Coverdale NS, Gati JS, Opalevych O, Perrotta A, Shoemaker JK. Cerebral blood flow velocity underestimates cerebral blood flow during modest hypercapnia and hypocapnia. J Appl Physiol (1985). 2014;117(10):1090–6. Epub 2014/07/12. doi: 10.1152/japplphysiol.00285.2014. PubMed PMID: 25012027.

45. Lucas SJE, Cotter JD, Brassard P, Bailey DM. High-intensity interval exercise and cerebrovascular health: curiosity, cause, and consequence. J Cerebr Blood F Met. 2015;35(6):902–11. doi: 10.1038/jcbfm.2015.49. PubMed PMID: WOS:000355575600003.

46. Hornby TG, Reisman DS, Ward IG, Scheets PL, Miller A, Haddad D, et al. Clinical Practice Guideline to Improve Locomotor Function Following Chronic Stroke, Incomplete Spinal Cord Injury, and Brain Injury. J Neurol Phys Ther. 2020;44(1):49–100. Epub 2019/12/14. doi: 10.1097/NPT.0000000000000303. PubMed PMID: 31834165.

47. Boyne P, Scholl V, Doren S, Carl D, Billinger SA, Reisman DS, et al. Locomotor training intensity after stroke: Effects of interval type and mode. Top Stroke Rehabil. 2020;27(7):483–93. Epub 2020/02/18. doi: 10.1080/10749357.2020.1728953. PubMed PMID: 32063178; PubMed Central PMCID: PMCPMC7429314.

